## Supplemental material for "Aetiology of Lobar Pneumonia Determined by Multiplex Molecular Analyses of Lung and Pleural Aspirate Specimens in The Gambia"

***Methods for multiplex PCR assay***

Total nucleic acid was extracted from a 200µl aliquot of lung and pleural aspirates (easyMAG, bioMériux, France) with an internal control. Extracts were subjected to quantitative multiplex PCR (Fast-track Diagnostics Resp-33 kit, Sliema, Malta) for a panel of 33 respiratory bacteria, fungi, and viruses with internal positive and negative control. The assay was structured in eight component multiplex sub-assays with three or four targets run on one plate. We used a Bio-Rad CFX96 thermocycler with programming as recommended by the manufacturer. Standard PCR curves were derived from plasmid standards during the testing to calculate pathogen load from cycle threshold values. The multiplex PCR included the following targets:

- *S. pneumoniae* (lytA),
- *Haemophilus influenze* sp. (ompP6),
- *H. influenzae* type b (bexA),
- *S. aureus* (shkv),
- *Chlamydia pneumoniae* (RNApbc),
- *Moraxella catarrhalis* (copB),
- *Klebsiella pneumoniae* (khe),
- *Legionella* sp. (16SrRNA),
- *Pneumocystis jirovecii* (mtlsurRNA),
- *Bordetella pertussis* (is481),
- *Salmonella* sp. (ttrB),
- Influenza A (pos1), B (seg8ns1nep) and C (mtx),
- Cytomegalovirus (us7&8),
- Parainfluenza virus 1 (hnmRNA), 2 (hnmRNA), 3 (hnmRNA) and 4 (fus),
- Rhinovirus (utr),
- Coronaviruses NL63 (ncpn), 229E (ncpn), OC43 (ncpn) and HKU1 (ncpn),
- Respiratory syncytial virus A (nucap) and B (numRNA),
- Metapneumovirus A (fuglF) and B (fuglF),
- Adenovirus (hex),
- Bocavirus (np1),
- Enterovirus (dom4&5),
- Parechovirus (utr),
- *Mycoplasma pneumoniae* (adP1),

Data were not used for *K. pneumoniae* and Legionella spp. Interpretation for some targets required combinations of results: if rhinovirus only was detected then the specimen was deemed rhinovirus positive, whereas if rhinovirus and enterovirus were detected then the specimen was deemed enterovirus positive; if *H. influenzae* type b and *H. influenzae* were detected the specimen was deemed positive for *H. influenzae* type b, whereas if *H. influenzae* only was detected the specimen was deemed positive for *H. influenzae* non-type b.

***Clinical characteristics of patients***

Table 1 in the manuscript describes the characteristics of the patients in three categories: no lobar consolidation, lobar consolidation and no lung/pleural aspirate, and lobar consolidation and lung/pleural aspirate. Compared to patients without lobar pneumonia, those with lobar pneumonia had greater respiratory rate (*p*<0.0001), lower oxygen saturation (*p*=0.034), and less wheeze (*p*<0.0001), whereas heart rate (*p*=0.59), temperature (*p*=0.73), prostration (*p*=0.25), weight-for-height z-score <-3 in young children (*p*=0.28) and severe underweight in older children and adults (*p*=0.36) were not significantly different. Respiratory rate (*p*=0.50), heart rate (*p*=0.20), temperature (*p*=0.12), prostration (*p*=0.20), weight-for-height z-score <-3 in young children (*p*=0.62), and severe underweight in older children and adults (*p*=0.86) were not significantly different in patients with lobar pneumonia who did or did not have a lung aspirate, although wheeze was more frequent in patients without lung aspirate (76/562 versus 11/181, *p*=0.007) and oxygen saturation was greater (*p*=0.017). Bacteremia was more likely in patients who had a lung aspirate (31/178, 17%) compared to those without a lung aspirate, irrespective of whether lobar pneumonia was present on chest radiograph (113/2119, 5%). Ninety-six patients died (3.8%) with similar proportions in the three clinical categories.

***Quantification of pathogen load***

The greatest pathogen load in lung specimens was associated with *S. pneumoniae* (median 5.34 [IQR 3.73, 6.24] log_10_ copies/ml), *H. influenzae* non-type b (median 6.07 [IQR 5.32, 6.86] log_10_ copies/ml) and parainfluenza virus (PIV) 1 (median 6.46 [IQR 4.74, 10.93] log_10_ copies/ml) positive specimens (Supplementary Table 1). Low pathogen load was associated with *S. aureus* (median 2.15 [IQR 1.68, 4.14] log_10_ copies/ml), bocavirus (median 2.77 [IQR 2.19, 3.40] log_10_ copies/ml]), and cytomegalovirus (2.57 [IQR 2.38, 3.71] log_10_ copies/ml) positive specimens.

**Supplementary table 1. Organism-specific quantification of pathogen load in 156 lung and 4 pleural aspirate specimens**

| **Organism** | **Quantification of organism**  **(median[IQR]; min, max); log_10_ copies per ml** |
| --- | --- |
| **Bacteria** |  |
| *Streptococcus pneumoniae* (n=68) | 5.34 (3.73 – 6.24); 1.44, 9.58 |
| *Staphylococcus aureus* (n=26) | 2.15 (1.68 – 4.14); 1.43, 8.49 |
| *Haemophilus influenzae* type b (n=11) | 4.18 (2.26 – 6.30); 1.56, 9.11 |
| *Moraxella catarrhalis* (n=8) | 4.40 (3.71 – 5.50); 2.63, 6.30 |
| *Salmonella* species (n=8) | 3.01 (1.74 – 5.29); 0.86, 9.07 |
| *Haemophilus influenzae* non-type b (n=6) | 6.07 (5.32 – 6.86); 4.88, 8.21 |
| *Bordetella pertussis* (n=4) | undef (undef); 0.30, 4.32 |
| *Chlamydia pneumonia* (n=3) | 3.60 (undef); 2.13, 4.73 |
| **Viruses** |  |
| Bocavirus (n=11) | 2.77 (2.19 – 3.40); 1.53, 4.76 |
| Parainfluenza 1 (n=8) | 6.46 (4.74 – 10.93); 4.32, 12.50 |
| Influenza C (n=7) | 4.47 (4.21 – 5.64); 3.72, 6.85 |
| Cytomegalovirus (n=6) | 2.57 (2.38 – 3.71); 1.45, 5.89 |
| Coronavirus HKU1 (n=4) | 3.93 (undef); 3.77, 4.46 |
| Coronavirus 43 (n=4) | 4.77 (undef); 4.25, 5.37 |
| Respiratory syncytial virus (n=3) | 6.59 (undef); 5.07, 7.07 |
| **Fungi** |  |
| *Pneumocystis jirovecii* (n=9) | 2.82 (2.52 – 3.37); 2.14, 7.42 |

Note: organisms listed were detected in three or more of 160 specimens. *B. pertussis* PCR Ct values were too great to allow quantification for three of seven specimens. The results of pathogen quantification in lobar pneumonia are subject to variation in the small volumes of specimen obtained and its dilution in 1ml of sterile saline.

***Effectiveness of PCV to prevent pneumococcal pneumonia***

**Supplementary table 2. Association of pneumococcal pneumonia with PCV vaccination status**

| **Pneumonia aetiology by culture of blood or lung/pleural aspirate** | **Number of PCV doses (PCV7 or PCV13)** | | **Total**  **N** | **Odds ratio**  **(95% CI)** |
| --- | --- | --- | --- | --- |
|  | ≥2 doses | 0 doses |  |  |
| Age 2-11 months | N=540 | N=184 |  |  |
| Culture pneumococcal | 5 | 5 | 10 |  |
| Culture non-pneumococcal | 535 | 179 | 714 | 0.33 (0.08, 1.47) |
| Proportion culture pneumococcal | 0.009 | 0.027 | 724 |  |
| Age 12-23 months | N=515 | N=81 |  |  |
| Culture pneumococcal | 15 | 2 | 17 |  |
| Culture non-pneumococcal | 500 | 79 | 560 | 1.19 (0.27, 10.9) |
| Proportion culture pneumococcal | 0.029 | 0.025 | 577 |  |
| Age 2-4 years | N=230 | N=218 |  |  |
| Culture pneumococcal | 9 | 15 | 24 |  |
| Culture non-pneumococcal | 221 | 203 | 424 | 0.55 (0.21, 1.38) |
| Proportion culture pneumococcal | 0.039 | 0.069 | 448 |  |
| Combined age strata 2-59 months, ^a^M-H age-stratified odds ratio = 0.57 (0.31, 1.06), ^b^*p*=0.076 | | | | |
| **Pneumonia aetiology by culture of blood or lung/pleural aspirate or PCR on lung/pleural aspirate** | | | | |
| Age 2-11 months | N=540 | N=184 |  |  |
| PCR or culture pneumococcal | 8 | 8 | 16 |  |
| Not PCR or culture pneumococcal | 532 | 176 | 684 | 0.33 (0.11, 1.03) |
| Proportion PCR or culture pneumococcal | 0.015 | 0.043 | 708 |  |
| Age 12-23 months | N=515 | N=81 |  |  |
| PCR or culture pneumococcal | 22 | 4 | 26 |  |
| Not PCR or culture pneumococcal | 493 | 77 | 570 | 0.86 (0.28, 3.52) |
| Proportion PCR or culture pneumococcal | 0.043 | 0.049 | 596 |  |
| Age 2-4 years | N=230 | N=218 |  |  |
| PCR or culture pneumococcal | 13 | 21 | 34 |  |
| Not PCR or culture pneumococcal | 217 | 197 | 414 | 0.56 (0.25, 1.21) |
| Proportion PCR or culture pneumococcal | 0.057 | 0.096 | 448 |  |
| Combined age strata 2-59 months, ^a^M-H age-stratified odds ratio = 0.54 (0.33, 0.90), ^b^*p*=0.017 | | | | |

^a^Mantel-Haenzel age-stratified odds ratio. ^b^Fisher’s exact *p*-value.
